# Utility of Pan-Family Assays for Rapid Viral Screening: *Reducing Delays in Public Health Responses During Pandemics*

**DOI:** 10.1101/2020.05.24.20112318

**Authors:** Michael Erlichster, Gursharan Chana, Daniela Zantomio, Benjamin Goudey, Efstratios Skafidas

## Abstract

**Background:** The SARS-CoV-2 pandemic has highlighted deficiencies in the testing capacity of many developed countries during the early stages of emerging pandemics. Here we describe the potential for pan-family viral assays to improve early accessibility of large-scale nucleic acid testing.

**Methods:** Coronaviruses and SARS-CoV-2 were used as a case-study for investigating the utility of pan-family viral assays during the early stages of a novel pandemic. Specificity of a pan-coronavirus (Pan-CoV) assay for viral detection was assessed using the frequency of common human coronavirus (HCoV) species in key populations. A reported Pan-CoV assay was assessed to determine sensitivity to SARS-CoV-2 and 59 other coronavirus species. The resilience of the primer target regions of this assay to mutation was assessed in 8893 high quality SARS-CoV-2 genomes to predict ongoing utility during pandemic progression.

**Findings:** Due to infection with common HCoV species, a Pan-CoV assay would return a false positive for as few as 1% of asymptomatic adults, but up to 30% of immunocompromised patients displaying symptoms of respiratory disease. Two of the four reported pan-coronavirus assays would have identified SARS-CoV-2 and we demonstrate that with small adjustments to the primers, these assays can accommodate novel variation observed in animal coronaviruses. The assay target region of one well established Pan-CoV assay is highly resistant to mutation compared to regions targeted by other widely applied SARS-CoV-2 RT-PCR assays.

**Interpretation:** Pan-family assays have the potential to greatly assist management of emerging public health emergencies through prioritization of high-resolution testing or isolation measures, despite limitations in test specificity due to cross-reactivity with common pathogens. Targeting highly conserved genomic regions make pan-family assays robust and resilient to mutation of a given virus. This approach may be applicable to other viral families and has utility as part of a strategic stockpile of tests maintained to better contain spread of novel diseases prior to the widespread availability of specific assays.

## Introduction

During the early stages of the severe acute respiratory syndrome coronavirus 2 (SARS-CoV-2) pandemic many countries exhibited an extreme shortage of SARS-CoV-2 nucleic acid test kits. This resulted in a weeks to months-long period where testing could only be performed in a limited capacity at select test centers, with a focus on symptomatic patients with a history of travel or association with a known case. The inability to perform extensive testing was particularly impactful in the current pandemic due to the greater than normal infectivity of asymptomatic patients^1^, placing a greater burden on molecular diagnostic tools for identifying and containing disease spread. The precise cause of this shortage has not yet been fully investigated but is likely a combination of the unprecedented global demand for test reagents and equipment, disruption to supply chains caused by the pandemic and regulatory restrictions limiting the ability of some nations to expand test capacity^2^. These shortcomings highlight that the current testing infrastructure and capacity expansion strategies are not rapid enough to counter disease spread during the early stages of some pandemics.

In response to SARS-CoV-2, governments will likely invest in a more extensive and agile network of testing equipment, stockpile test reagents and consumables and streamline test certification protocols. While this will greatly enhance the speed at which testing capacity can be increased, there will still be a vital period between the time a novel pathogen emerges and the time that tests are widely available at high capacity, which can hamper efforts to contain disease spread. A strategy which may allow wide-spread testing of novel pathogens in a more-timely manner is the use of pre-emptively developed pan-family assays: molecular diagnostic tests targeted at a family of viruses rather than a single species. Pan-family assays targeting several viral families have been developed for research applications and are routinely used during pathogen characterization and in retrospective epidemiological studies^3,4^.

To reduce the likelihood of severe test shortages in future pandemics, we propose a proactive strategy involving the large-scale stockpiling of pan-family targeted PCR diagnostic kits for use during the early stages of viral pandemics **(Figure 1)**. In this strategy, pan-family tests are developed, certified and stockpiled in large volume prior to disease emergence. Following confirmation of test applicability to the novel pathogen, it may then be applied to deliver unrestricted testing while a species-specific test is produced.

**Figure 1.**
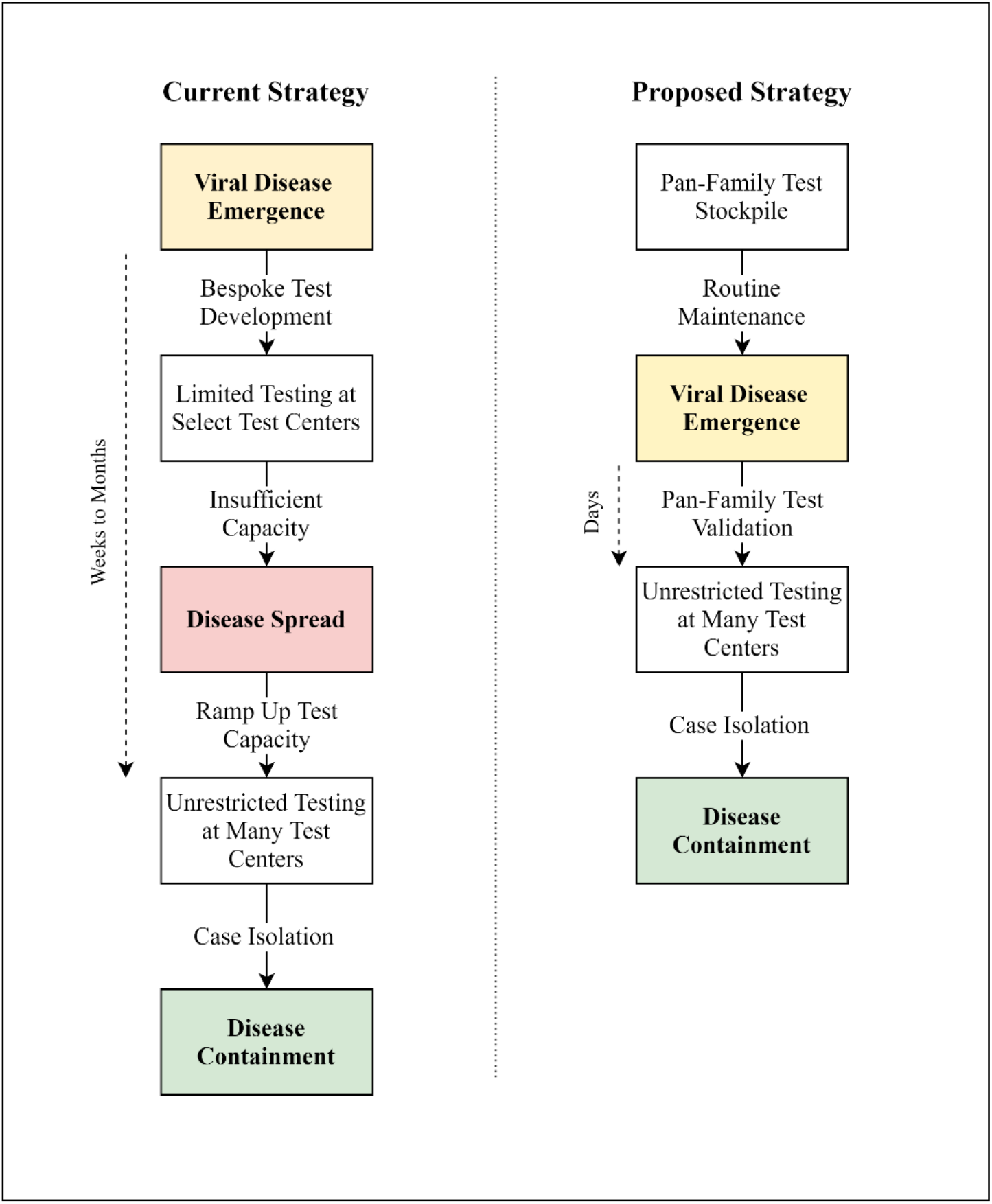
Current and proposed viral disease testing strategies. In the current strategy a delay of weeks to months is observed between disease emergence and unrestricted testing, allowing a disease to spread. By using pan-family tests widespread testing is available in a shortened timeframe and disease spread may be contained.

Here we assess, using coronaviruses and SARS-CoV-2 as a case study, whether pan-family targeted assays are a practical tool for high-throughput screening of infection during the early stages of a pandemic. We consider the frequency of common viral species, the capability of reported pan-family assays to accommodate observed variation and the mutation rate of the pan-family assay target regions. Given the significant global economic and social disruption caused by the SARS-CoV-2 pandemic, we argue that despite some limitations in the specificity and sensitivity of pan-family assays, a strategic stockpile is prudent as a first-line mass-screening technique.

## Methods

### Review of common HCoV epidemiolocal characteristics

A search using the terms “HCoV” “epidemiology” and “asymptomatic” was performed in PubMed to identify studies describing HCoV infection characteristics. Studies highlighting the following aspects of HCoV infection were selected and summarized; frequency in children or the elderly, frequency in asymptomatic individuals, frequency in large study cohorts and frequency during local epidemics.

### Validation and modification of a reported Pan-CoV assays

Coronavirus-family reference sequences were identified and downloaded from the NCBI Virus database^5^. Sequences for a total of 60 species were identified **(Supplementary Table 1)**, including the 7 species known to infect humans (HCoV-229E, HCoV-NL63, HCoV-HKU1 and HCoV-OC43, SARS-CoV, MERS-CoV and SARS-CoV-2). Pan-coronavirus assay primer targets were derived from four reported assays^6-9^ and aligned with the SARS-CoV-2 reference genome (NC_045512) to assess consensus between sequences using the Clustal Omega Multiple Sequence Alignment tool^10^.

The nucleic acid sequence and predicted protein sequence for each species was aligned and assessed for identity with Pan-CoV primer targets using the software package Geneious Prime 2020.1.2 (https://www.geneious.com). Primers were modified to accommodate observed nucleic acid variation as well as nucleic acid sequences expected from the observed protein variation.

### Assessment of the Pan-CoV primer target site mutation rate

Mutation frequency of the Pan-CoV target site was assessed using 8,899 high coverage (<1% N calls and <0.05% unique amino acid mutations) full length (>29,000 base pairs) SARS-CoV-2 genomes downloaded from the GISAID EpiCoV database^11^. Analysis was performed with the R Biostrings package^12^ to identify mismatches in the primer target sites. Three samples were excluded due to missing sequence data in the primer target region.

## Results

### Specificity of a Pan-CoV assay to a novel pathogen - impact of endemic coronavirus species

A key factor in the utility of pan-family assays is the frequency of common species of the target family in the general population, as these will interfere with the specificity of an assay for the novel pathogen. For the coronavirus family there are 4 endemic and common species that infect humans (HCoV-229E, HCoV -NL63, HCoV-HKU1 and HCoV-OC43), typically causing mild disease but can lead to severe or fatal infections in frail or immunosuppressed patients. It remains to be seen whether the current SARS-CoV-2 pandemic will persist at low levels in the general population following disease control through social distancing, therapeutic measures or acquisition of herd immunity^13^. Non-endemic zoonotic coronavirus species (SARS-CoV, MERS-CoV), while highly pathogenic, would not affect the specificity of a Pan-CoV assay for a novel pathogen as they are not observed at a relevant frequency. However, Pan-CoV assay sensitivity to these pathogens is important as they are informative of species with zoonotic potential.

**Table 1** summarizes studies reporting the prevalence of common HCoV infection. HCoV infections are typically detected in 2-10% of patients exhibiting acute respiratory illness (ARI), although during local HCoV epidemics frequency of infection may be as high as 30% in symptomatic patients. Additionally, HCoV infections display a seasonality, typically observed in winter months. Children, both experiencing ARI and asymptomatic have a high occurrence of HCoV infection (4-10%). In asymptomatic adults HCoV infection is less well studied, but reported values are lower at ~1%. Studies of seroprevalence of HCoV antibodies indicate that virtually all individuals have had prior exposure to at least one form of coronavirus, with first exposure common in childhood^14^. This high frequency of childhood infection may explain the lower prevalence of asymptomatic infection in adults due to a partial acquired immunity^15^, though limited data exists describing the immunizing effect of HCoV infection^16^.

**Table 1:** Population Frequency of Common HCoV Infection in Highlighted Studies. ARI – Acute Respiratory Illness. ARTI – Acute Respiratory Throat Infection

| Study | Population | Observed HCoV infection rates | Ref. |
| --- | --- | --- | --- |
| Van der Zalm (2009) | 230 respiratory samples routinely collected from 19 children across 6 months | Overall – 8.7%<br>Asymptomatic – 7.7%<br>Symptomatic Samples – 9.1% | 17 |
| Gaunt (2010) | 11,661 respiratory samples from 7,383 patients as part of routine respiratory virus screening. | Overall – 2.3%<br>Asymptomatic - 1.08%<br>7-12 months old – 4.86% | 18 |
| Prill (2012) | 1481 hospitalized children with ACI or fever and 742 controls. | Overall – 7.5%<br>Hospitalized – 7.6%<br>Asymptomatic – 7.1% | 19 |
| Cabeca (2013) | 1087 respiratory samples from patients with ARI. 50 Asymptomatic controls. | Overall – 7.7%<br>Asymptomatic Adults – 0.0%<br>All ARI patients - 8.1%<br>Patients with Comorbidities – 12.4% | 20 |
| Lepiller (2013) | 6,014 respiratory samples collected for routine viral diagnostics. | Overall – 5.9%<br>Immunosuppressed Patients– 6.7%<br>During Local Epidemic – ~16% | 21 |
| Matoba (2015) | 4,342 respiratory samples from pediatric patients with ARTI | Overall – 7.6%<br>During Local Epidemic – 28.5% | 22 |
| Zhang (2015) | Meta-analysis of 489,651 patients with ARTI | Overall- 2.6% | 23 |
| Yip (2016) | 8275 respiratory samples from patents with ARTI | Overall – 0.9%<br>0-10 years old – 0.9%<br>Over 80 years old - 1.5% | 24 |
| Liu (2017) | 3,298 respiratory samples from pediatric patients with ARI | Overall – 2.37%<br>During Local Epidemic – 10% | 25 |
| Zeng (2017) | 11,399 respiratory samples from hospitalized pediatric patients with ARTI | Overall – 4.3%<br>7-12 months old – 5.9% | 26 |
| Killerby (2018) | 854,575 HCoV tests from 117 Laboratories as part of routine respiratory virus screening. | Overall – 4.6%<br>During Local Epidemic – 12.4% | 27 |
| Heimdal (2019) | 3,458 samples from hospitalized pediatric patients with ARTI and 373 samples from controls. | Overall – 9.1%<br>Asymptomatic controls – 10.2%<br>Hospitalized – 9.1% | 28 |

Considering the observed frequencies of common coronavirus species **(Table 1)**, a Pan-CoV assay would be expected to be sufficiently specific for a novel pathogen to allow for broad population screening, wherein a positive test is used to guide infection controls such as self-isolation. The screening specificity is strongest in asymptomatic adults where only ~1% of the population would be expected to return a false positive due to HCoV infection. Greater caution must be taken in populations with a higher HCoV infection rate such as children, symptomatic patients with comorbidities, or populations experiencing a local HCoV epidemic. Due to relatively high frequency of common HCoV infection in these populations additional follow up testing may be necessary to improve the utility to public health officials.

### Sensitivity of Pan-CoV assays to a novel pathogen – performance of reported assays on observed coronavirus species from animal populations

Several consensus reverse-transcription PCR (RT-PCR) assays targeting the highly conserved coronavirus RNA-dependent RNA polymerase gene with a pool of degenerate primers have been previously described for research applications^6-9^. These assays have been used to identify and analyze previously unknown coronavirus species^3^ and perform retrospective analysis of clinical samples to assess disease prevalence^20^.

**Figure 2** compares the target sequences of reported Pan-CoV assays with the SARS-CoV-2 reference genome. Only 2 of the 4 reported assays tested, the Moes/Vijgen (2005) update of the Stephensen (1999) assay and the recent Hu (2018) assay, accommodate the SARS-CoV-2 genome without any primer-template mismatches. While these mismatches may not be sufficient to prevent detection, they are undesirable as they may reduce reaction sensitivity^29^. This result highlights the importance of routine maintenance of pan-family assays, ensuing degenerate primers accommodate annotated variation from animal populations.

**Figure 2:**
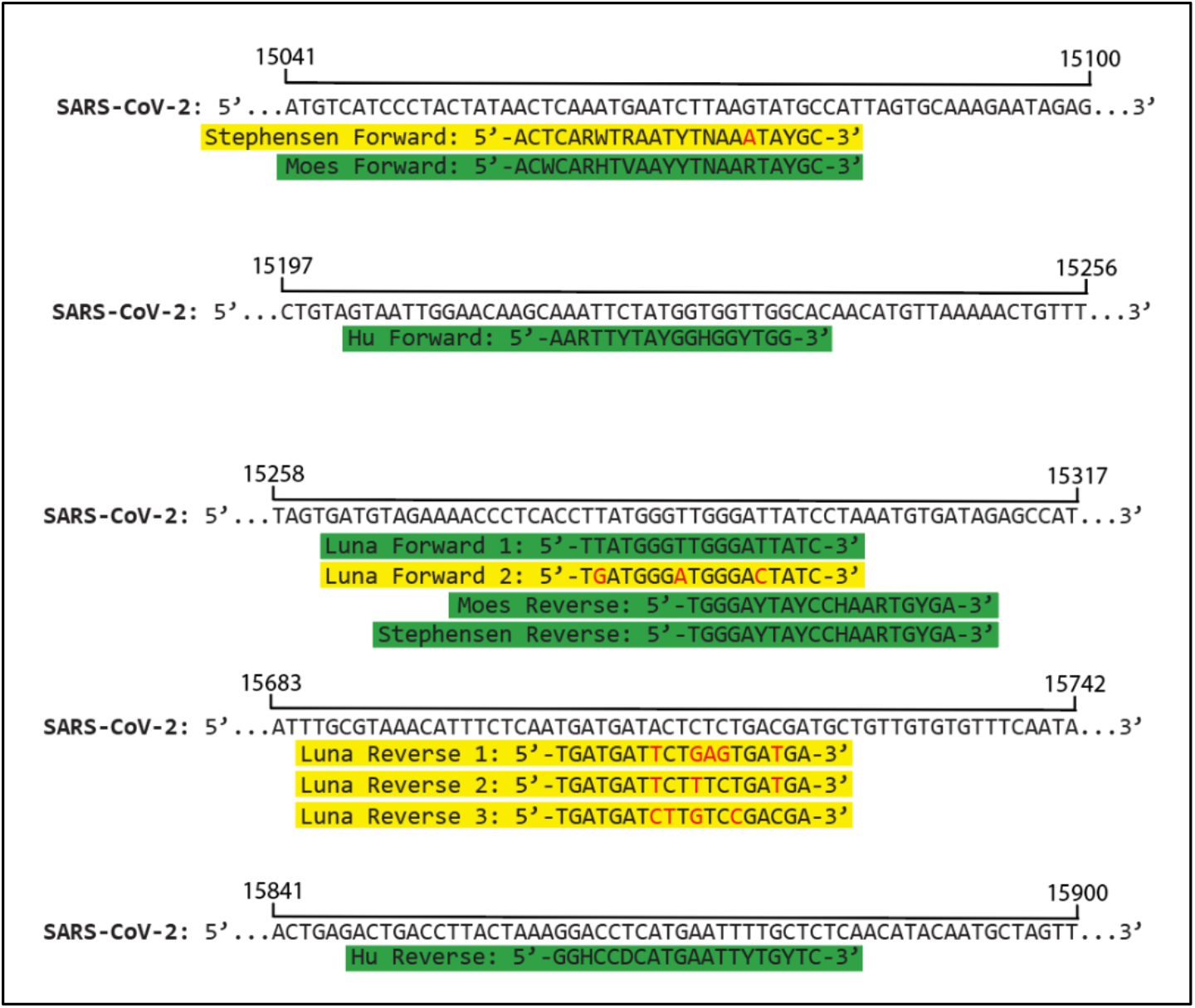
Alignment of 4 Reported Pan-Coronavirus Primer Targets and the SARS-CoV-2 Genome. IUPAC nucleotide code used where degenerate primers are reported. Each of the 5 rows shows a different region of the SARS2 genome. Primers with mismatches between target sites and SARS-CoV-2 genome are highlighted in yellow, with mismatches in red. Primers targets with a 100% match to the SARS-CoV-2 genome highlighted in green.

In contrast to the other reported Pan-COV assays, the Moes/Vijgen assay (MPC) has a detailed protocol for analysis of respiratory pathogens in human samples^7^. To better predict the sensitivity of this assay to novel pathogens, we compared the MPC primer sequences to the respective primer target sites in the genomes of 60 coronavirus species **(supplementary Table 2)**. The majority of these species were reported following the development of the MPC assay, and as such are informative of the capacity of existing tests to detect novel pathogens.

Of the 60 coronavirus reference sequences investigated, 20 contained a mismatch between the MPC degenerate primer target and the viral genome, suggesting the MPC assay would be sub-optimal for detection of these species **(Figure 3, Supplementary Table 1)**. Of these 20 species, only 4 contained a genomic variant resulting in a change to the amino acid motifs encoded by the target region. **(Supplementary Table 2)**. This highlights the importance of accommodating unobserved but likely variation, such as silent mutations, when designing pan-family assays. Indeed, degenerate primers designed given the observed protein motifs of these 60 species **(Supplementary Tables 3)** are identical to those designed from the observed sequence data **(Supplementary Table 4)**.

**Figure 3:**
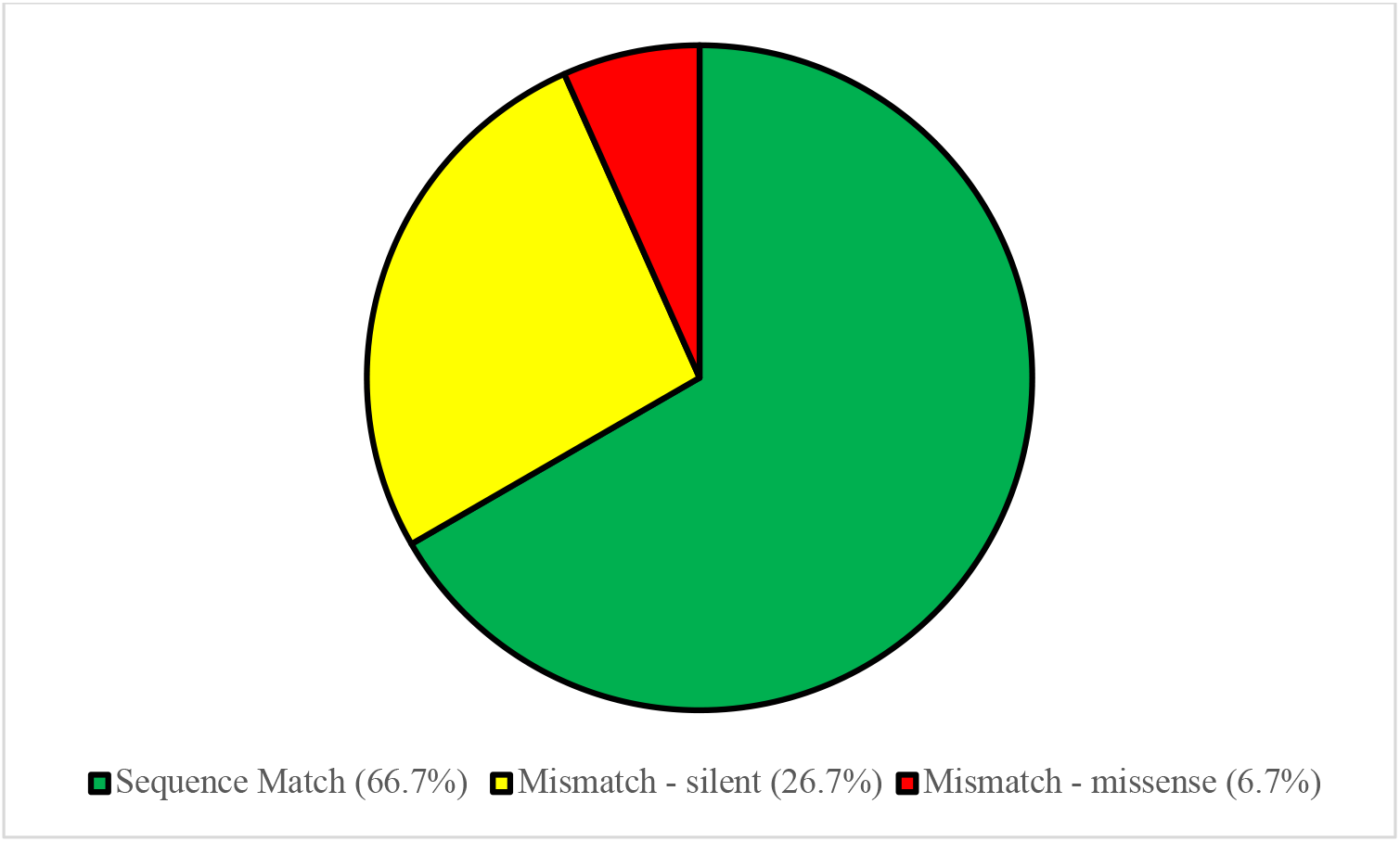
Sensitivity of the MPC Pan-CoV assay to coronavirus species observed in animal populations. A complete sequence match between the MPC primer target and the genomic sequence is observed for 40/60 (66.7%) of species, with a mismatch caused by a silent mutation observed in 16/60 (26.7%)of species and a mismatch resulting in an amino acid change observed in the remaining 4/60 (6.7%) of species.

### Resistance of the MPC target region to mutation

Another application of a robust pan-family assay is as a fallback to protect against reduction in test sensitivity of species-specific assays due to mutation of the viral pathogen during pandemic progression. To assess the resilience of the MPC assay to mutation-derived errors, we assessed mutation frequency of the MPC target sites in 8,893 high-coverage SARS-CoV-2 genomes. Only a single sample with a mutation in the MPC target region was observed, EPI_ISL_414596, translating to an observed mutation rate of 0.01%. This is an order of magnitude lower than the observed mutation rate for the CDC primer regions (0.4-2.58%) and the China CDC primer regions (0.74-16.19%) in the same dataset as reported on the GISAID platform^11^. These results indicate that the MPC target region is robust to mutation and may provide ongoing value in the case of emergence of a viral strain with mutations in the target regions of high specificity assays.

## Discussion

Nucleic acid testing for the identification of infected individuals is one the most valuable tools for controlling pathogen spread, especially for diseases with asymptomatic carriers. The recent SARS-CoV-2 pandemic has revealed deficiencies in the testing capacity of many countries, resulting in a period where testing was highly restricted, preventing optimal disease containment. The use of family-wide viral assays may reduce the time before mass testing is available by allowing tests to be developed and distributed before pathogen emergence. Here we demonstrate that a Pan-CoV test can be an effective tool for management of novel coronavirus pathogens. These results may also be more broadly applicable for the implementation of pan-family assays for the detection of pathogens from other viral families. **Table 2** summarizes the advantages and disadvantages of a Pan-family assay compared with the current high-specificity testing standard approach.

**Table 2:** Advantages and disadvantages of a Pan-family assay compared with current high-specificity approaches.

|  | **Advantages** | **Disadvantages** |
| --- | --- | --- |
| **Specific-Virus Assay** | - High specificity provides confident diagnostic value.<br>- Target optimization allows for detection of very low vial load.<br>- Well validated standard operating procedures for core technology. | - Lack of availability during early stages of a pandemic.<br>- Rush to develop and distribute in response to a novel pathogen may result in quality control issues. |
| **Pan-Family Assay** | - Stockpiling prior to pandemic development enables rapid and extensive population screening.<br>- Validated and quality controlled prior to pandemic development<br>- Can prioritize individuals for testing with short-supply Specific Virus Assays. | - Susceptible to off-target positives due to cross-reactivity with common pathogens, reducing diagnostic value or mandating follow up testing.<br>- Sensitivity and specificity of testing dependent on viral family and population of interest. |

Coronaviruses are a useful framework for assessing the utility of pan-family testing as several human coronaviruses are common globally and would interfere with specific identification of a novel coronavirus pathogen. However, even without any further testing a Pan-CoV test still allows for a large majority (>90%) of individuals, particularly asymptomatic adults, to avoid self-isolation measures with a negative test, though greater caution must be taken in populations with a higher HCoV infection rate such as children, symptomatic patients with comorbidities, or populations experiencing a local HCoV epidemic. This limitation can be mitigated through multiple strategies, most simply application of this tool as a first-line screen prior to confirmatory analysis with a more specific test. This would allow short-supply specific tests to be reserved as a confirmatory, rather than a primary tool, enabling a higher-throughput screen of large populations at ports-of-entry or for contact tracing during the early stages of pandemic. An alternative strategy, not reliant on the presence of a specific test, would be the addition of a second exclusionary reaction targeting known common species^30^, though this may not be an effective tool where a novel pathogen is highly genetically similar to a common species and may be overly burdensome for high-throughput population screening.

During the initial roll-out of SARS-CoV-2 testing kits by the CDC, several laboratories experienced disruptions due to faulty testing kits^31^. Pan-family assays, developed prior to an outbreak, would have a relaxed timeframe for test development, ensuring best-practice validation, quality control procedures, regulatory certification and laboratory accreditation can be achieved. Pan-family assays may also have additional ongoing utility as a fallback in the case of quality control issues or disease mutations compromising the accuracy of specific assay detection^32^. By targeting highly conserved regions and accommodating silent mutations, pan-family assays can be expected to be less prone to mutation induced errors, though routine comparison of primer target sites with observed viral strains is prudent. Further consideration must be given to the appropriate number of targets contained in a pan-family test to increase the likelihood that a novel pathogen is captured by an established assay, as well as the appropriate validation processes which should be undertaken following pathogen emergence before the approved use of a pan-family assay.

Our analysis has highlighted several limitations of reported pan-family tests resulting from an over-optimization on observed sequence variation rather than pre-emptive variation predicted from highly conserved protein motifs. Indeed, with the target regions applied by the MPC assay all possible genomic combinations translating to observed protein motifs are observed in the relatively small panel of reference coronaviruses. Broadening of primer degeneracy with the strategies described in this work, as well as routine comparison with novel animal viruses may assist in ensuring ongoing effectiveness of pan-family assays. However, highly degenerate primers, such as the modified primers described in this work, are likely to have reduced test sensitivity to viral load. This may be partially counteracted through the use of universal base analogues such as inosine or 5-nitroindole^33^ to reduced degeneracy or avoided through more judicious targeting of alternative highly conserved genomic motifs^9^. An alternative strategy may be the use of more relaxed reaction conditions to accommodate minor primer mismatches, though this may further reduce assay specificity^29^. These limitations must be appropriately characterized as part of pan-family assay development if to be applied in a clinical context.

A panel of pan-family assays would be required to provide a more robust tool for combating novel pathogens, with priority given to viral families associated with previous epidemics. Pan-family assays have been described for multiple viral families associated with recent epidemics, including filoviruses (Ebola Virus)^34^ and flaviviruses (Zika Virus, Dengue Virus, Yellow Fever Virus)^35^, and a similar analysis to the one presented here should be performed to assess the specificity and sensitivity of these assays for novel pathogens within the context of common viral species. Consistency in reaction conditions between pan-family assays or multiplexing of several assays into a single kit may further simplify the application of these tools during emerging pandemics.

A cost-effectiveness analysis of stockpiling pan-family assays is beyond the scope of this work given the novelty of the proposed testing strategies, and the unpredictability of pandemic emergence and characteristics. Most components required to run pan-family assays (nucleic acid-extraction kits, enzymes and buffers) may already be already sufficiently stocked by diagnostic laboratories or may be able to be repurposed for routine testing or research applications towards the end of their shelf life. The only unique components required are the specific test primers, which depending on oligonucleotide modifications, number of reactions and production volume can be expected to cost below 0.5USD/test, with a shelf life of at least 2 years. However, to increase the simplicity and rapidity of testing it may be practical to stockpile all reagents required to run a pan-family assay as part of a self-contained test-kit, despite the additional cost.

Strategic stockpiling of pan-family viral assays is a proactive alternative to current viral disease test strategies which may expedite testing during emerging pandemics. These assays have the potential to greatly assist management of emerging public health emergencies through prioritization of high-resolution testing or isolation measures, despite limitations in test specificity due to cross-reactivity with common pathogens. Extensive further development, validation and certification of pan-family assays is needed prior to application in broad clinical contexts. With appropriate design these tools may allow informative and high-throughput screening of millions of individuals within days of pathogen emergence.

## Data Availability

All data used in this manuscript is publicly available in in NCBI and GISAID databases as described in the methods section of the manuscript.

## Supplementary Table

**Supplementary Table 1.**
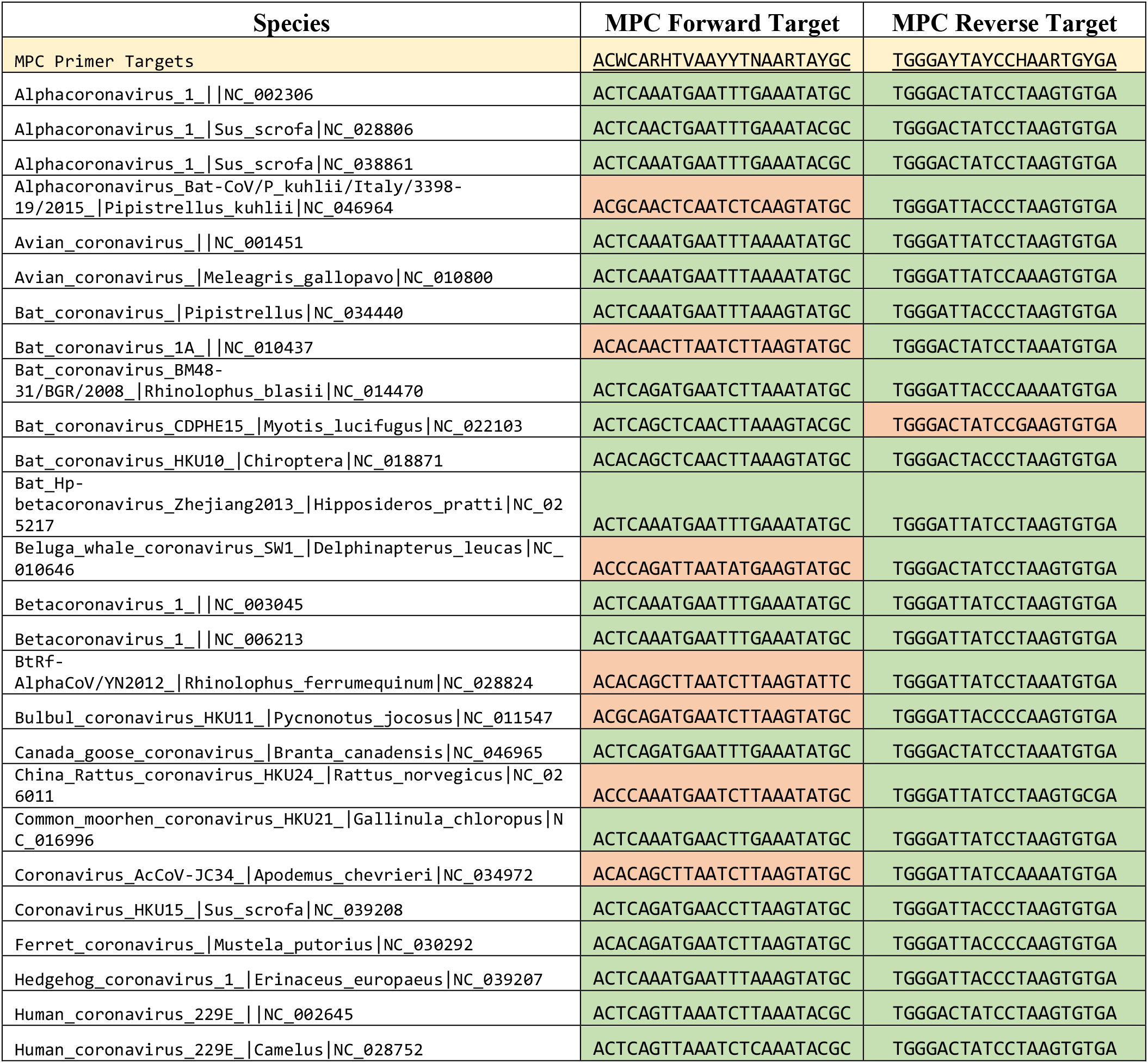
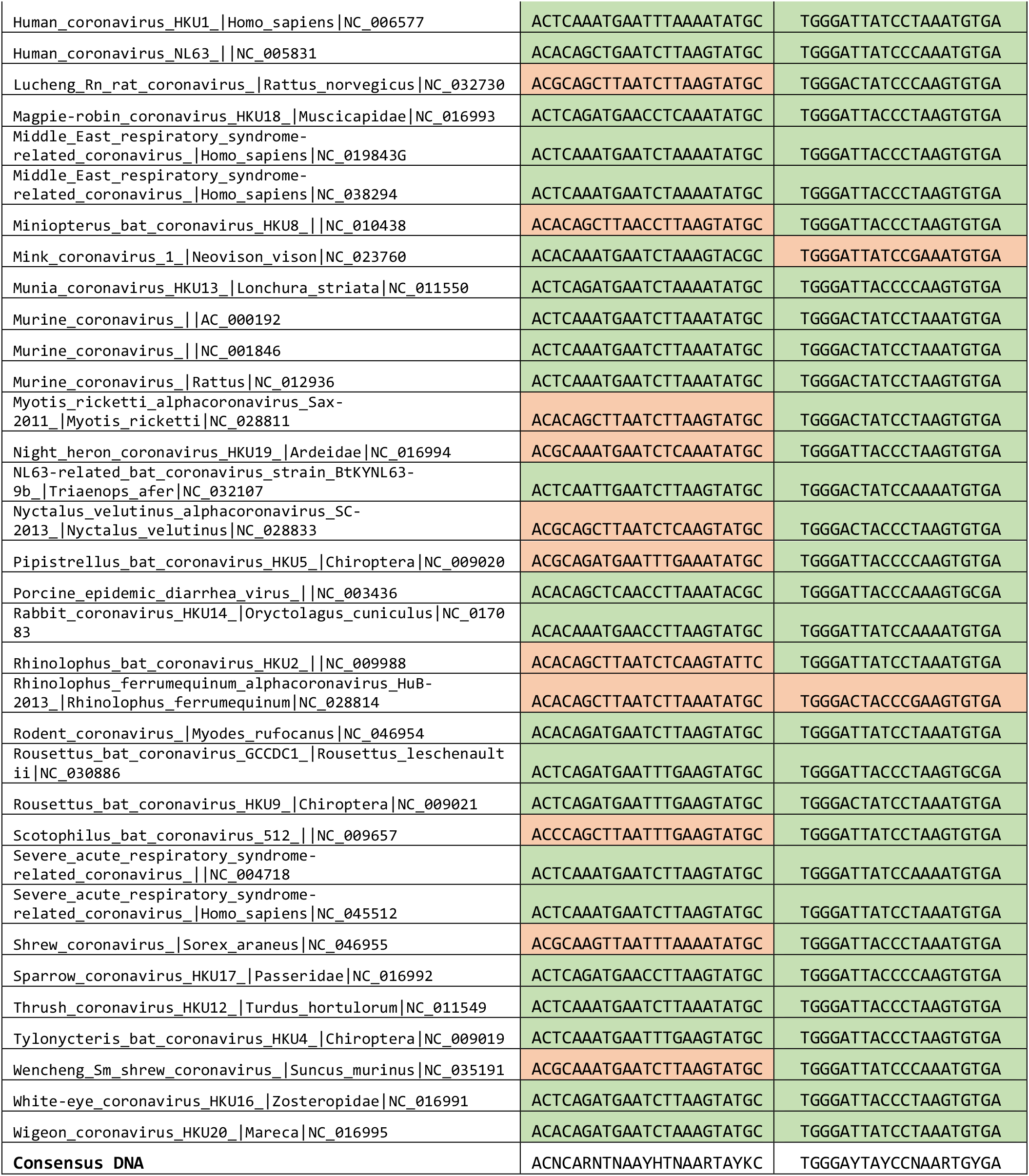
60 Coronavirus-family reference sequences used in this study and the nucleic acid sequence observed the MPC Primer Target Sites^7^. Genomic sequences which match the primer target sequences are highlighted in green while those which contain a variant not matching the primer target sequences are highlighted in orange. A consensus sequence for all observed variation is shown at the bottom of the table.

**Supplementary Table 2.**
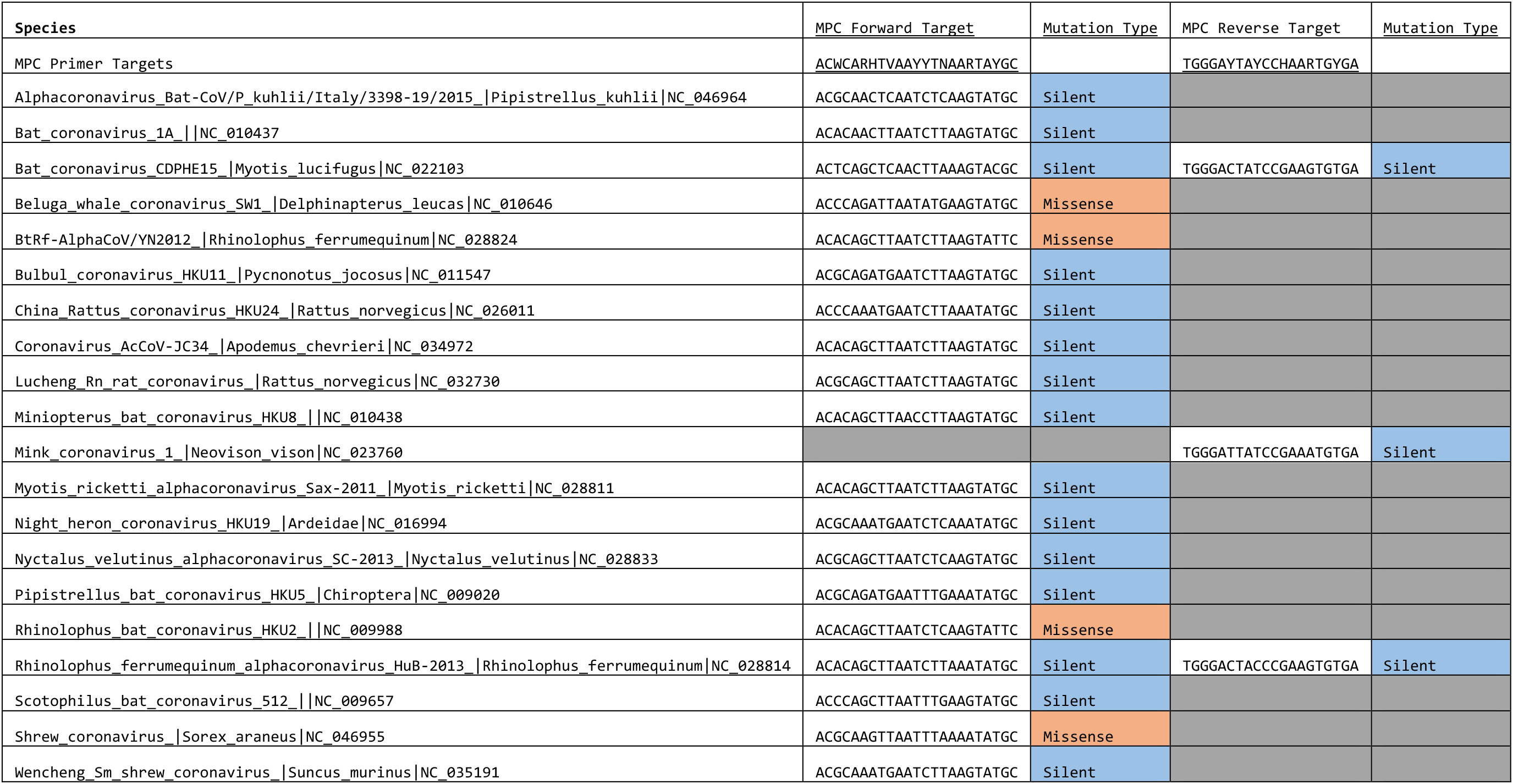
20 Coronavirus species with novel variation in the MPC Target sites and the effect of this variation on the protein motif encoded by this site. Silent mutations are highlighted in blue while missense mutations are highlighted in orange.

**Supplementary Table 3.**
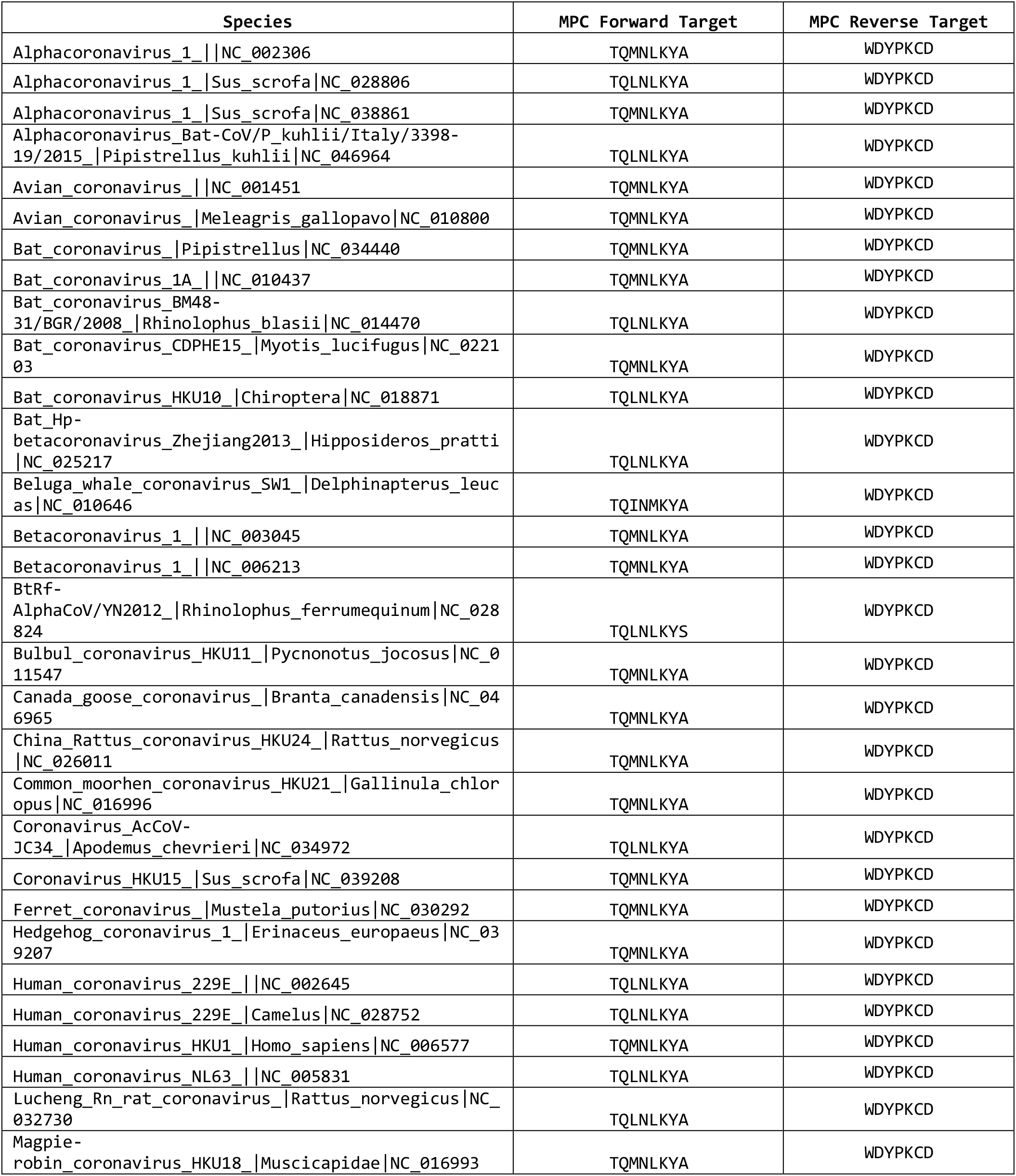
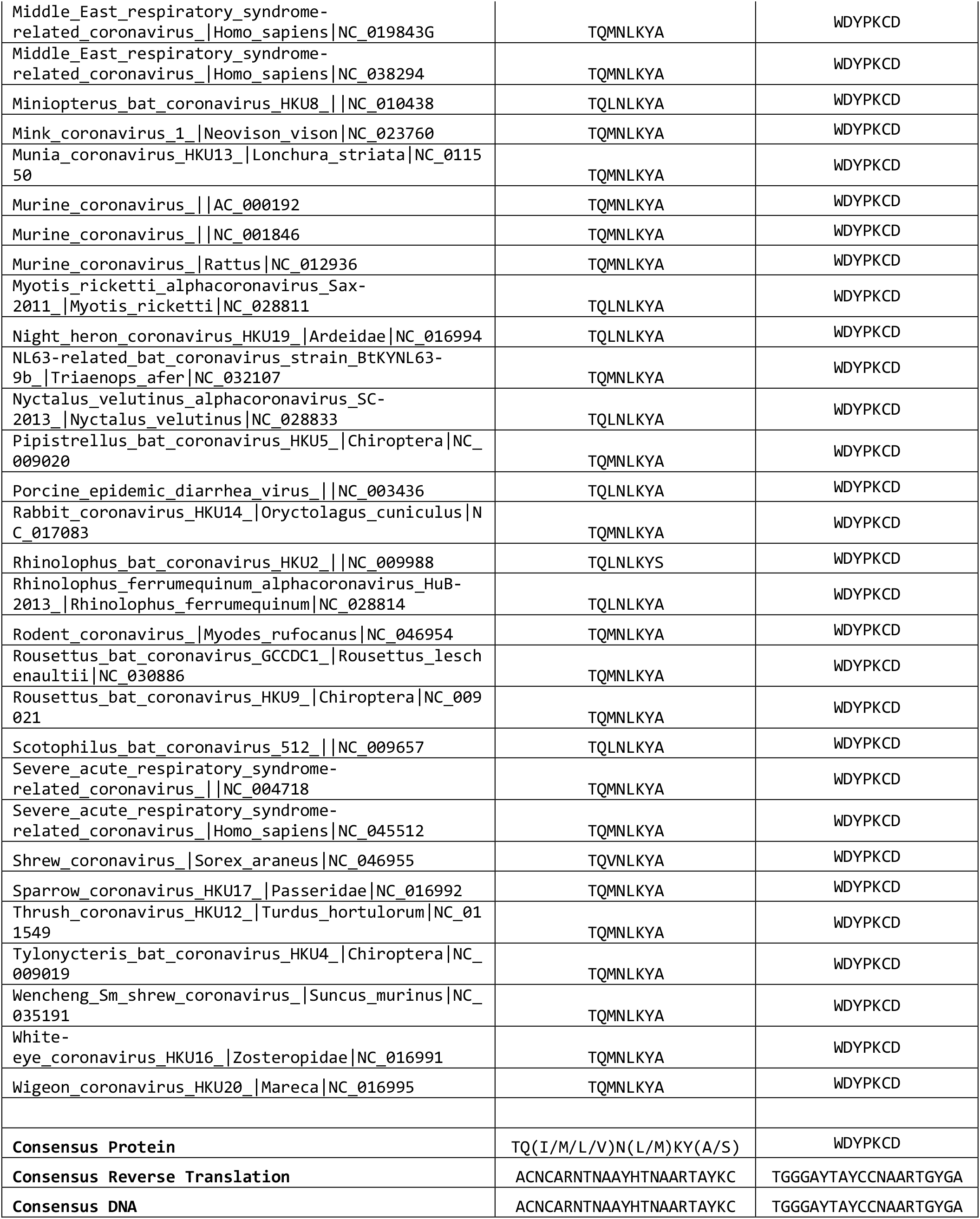
60 Coronavirus-family reference sequences used in this study and the protein sequence encoded by the MPC Assay Primer Target Sites^7^. A protein and nucleic acid consensus sequence for all observed variation is shown at the bottom of the table.

**Supplementary Table 4:**
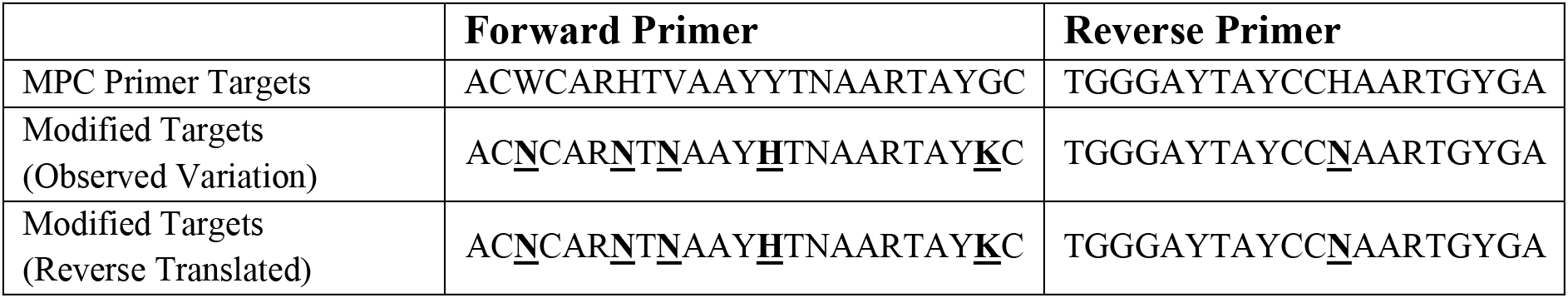
Revised Pan-Coronavirus primers accommodating observed nucleic acid and protein sequences. Modified Targets (Observed Variation) describe the degenerate primer sequences required to accommodate all observed genomic variation **(supplementary table 1)**. Modified Targets (Reverse Translated) describe a degenerate primer target sequence designed using the reverse translation of the observed protein sequences encoding the primer target sites **(supplementary table 3)**.

## Notes

### Competing Interest Statement

The authors have declared no competing interest.

### Funding Statement

No external funding received.

### Author Declarations

Only public and anonymised data from NCBI and GISAID databases was used in this study.

